# Self-Supervised Learning Can Distinguish Myelodysplastic Neoplasms from Clinical Mimics Using Bone Marrow Biopsies

**DOI:** 10.1101/2025.02.17.25322075

**Authors:** Vahid Mehrtash, Hortense Le, Bita Jafarzadeh, Sanam Loghavi, Guillermo Garcia-Manero, Aristotelis Tsirigos, Christopher Y. Park

**Affiliations:** Department of Pathology, NYU Langone Grossman School of Medicine, New York, New York; Department of Hematopathology, University of Texas, MD Anderson Cancer Center, Houston; Department of Leukemia, University of Texas, MD Anderson Cancer Center, Houston; Applied Bioinformatics Laboratories, New York University School of Medicine, New York, New York; Division of Precision Medicine, Department of Medicine, New York University School of Medicine, New York, New York

## Abstract

The diagnosis of myelodysplastic neoplasms (MDS) requires examination of the bone marrow for morphologic evidence of dysplasia. We sought to determine if a self-supervised learning (SSL) AI image analysis approach may be utilized to reliably distinguish MDS from its clinically relevant mimics using bone marrow biopsies (BMBx). Whole slide images (WSIs) of H&E- and reticulin-stained BMBx sections from 243 unique patients (89 MDS, 55 non-MDS cytopenic controls [NMCC], and 99 negative control [NC] cases) were segmented into tiles and analyzed. These tiles were then processed using the Barlow Twins SSL model to generate histomorphologic phenotype clusters (HPCs). Review of the HPCs revealed the clusters enriched in MDS captured known histopathologic features of MDS including hypercellularity, dysplastic and clustered megakaryocytes, increased immature hematopoietic cells, increased vascularity, fibrosis, and cell streaming patterns. Assessment of 95 MDS BMBx images from a second institution showed consistent HPC enrichment patterns, validating the robustness of the model. The trained ensemble model using H&E- and reticulin-stained slides distinguished MDS from NCs with an AUC of 0.89, and from age-matched, NMCCs with an AUC of 0.84. These findings demonstrate the potential of SSL approaches to capture diagnostically relevant morphologic patterns and to improve the reproducibility of MDS diagnosis.

## Introduction

Myelodysplastic neoplasms (MDS) are clonal hematopoietic disorders characterized by ineffective hematopoiesis, peripheral blood cytopenia(s), and dysplastic cells in the marrow.^1,2^ Diagnosis requires an integrated approach combining morphologic, genetic, and cytogenetic features.^3^ Identification of dysplastic morphologic changes, a key diagnostic criterion, is associated with significant inter- and intra-observer variability^4-7^. Given that morphologic assessment for dysplasia is required to establish a diagnosis of MDS and to distinguish MDS from benign conditions with similar presentations, this remains a significant potential diagnostic pitfall. Typically, evaluation for dysplasia is performed by assessing bone marrow aspirate (BMA) smears, since, in contrast to bone marrow biopsies (BMBx), a detailed cytologic examination of all three major lineages (erythroid, granulocytic, megakaryocytic) can be made.^8,9^ However, bone marrow aspirates are sometimes inadequate for evaluation due to the absence of marrow spicules and suboptimal preparation.^10,11^ A previous study demonstrated that supervised, deep learning-based morphological analysis of H&E stained BMBx from MDS patients with established diagnosis could predict genetic subtypes of MDS.^12^ However, these studies did not rigorously investigate the ability of AI-driven approaches to distinguish MDS from its clinical mimics based solely on BMBx images. In this study, we applied a self-supervised learning (SSL) image analysis approach to develop an algorithm capable of reliably discriminating MDS from its clinically relevant mimics. The model captured morphologic features required for the diagnosis of MDS, and thus may improve reproducibility and accuracy of diagnosis, particularly as AI-assisted algorithms gain broader adoption in hematopathology. Overall, these findings demonstrate that SSL can uncover domain-specific features in unlabeled data that reduce dependence on expert annotations.

## Methods

Details on case selection criteria and the analysis pipeline are provided in the supplementary file (Supplementary File).

Briefly, a total of 243 BMBx were included in the study, representing 89 diagnostic MDS cases based on established diagnostic criteria ^8^, 55 cytopenic cases with suspicion of MDS but resulting in non-MDS diagnoses (“non-MDS cytopenic controls” [NMCC]), and 99 unremarkable lymphoma staging bone marrow cases from non-MDS patients selected as negative control cases (“negative control” [NC]) (Fig.1A). In this study, both H&E- and reticulin-stained BMBx sections were included. To maximize representativeness, two slides per patient (with two levels per slide) were analyzed.

**Fig 1.**
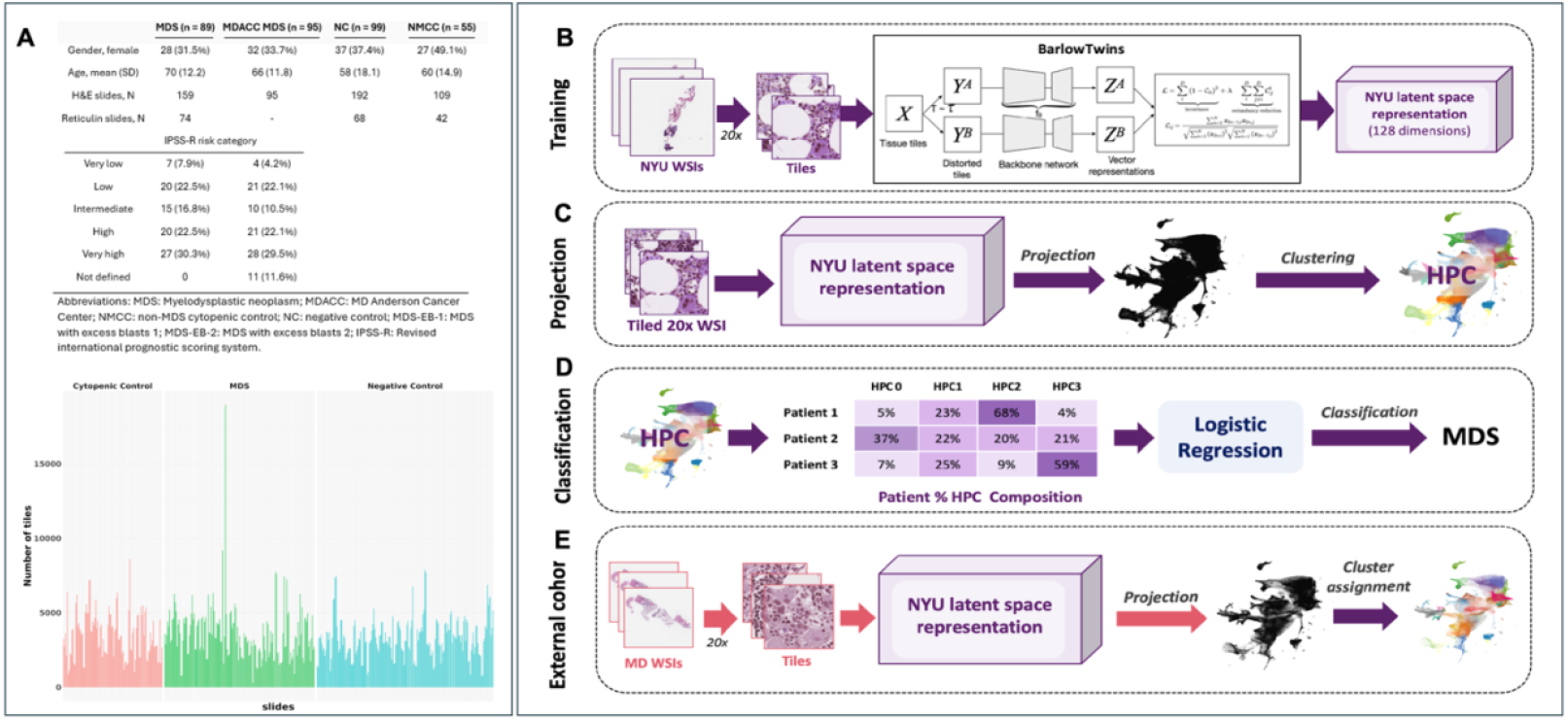
Pipeline overview for histomorphological phenotype learning (HPL) framework architecture. **(A)** Summary of patient samples and corresponding tile counts; **(B) Training phase**: whole slide images (WSI) are tiled at 20× magnification, stain normalized, and processed through a self-supervised learning framework, resulting in a 128-dimensional latent space representation for each tile. **(C-D) Projection and classification:** Following the dimensionality reduction and projection of latent space representations derived from the frozen backbone network, histomorphological phenotype clusters (HPC) are identified using the Leiden community detection algorithm on the resulting low-dimensional data. Subsequently, WSls are quantified in terms of the composition of various HPCs, providing a vectorized representation that can be used for training a regression model **(E) External validation**: The resulting HPCs were validated using an external cohort of MDS cases and the clusters were assigned to an unseen new dataset.

Whole slide images (WSIs) acquired at 20x magnification were segmented into 224×224-pixel tiles using DeepPATH.^13^ We utilized the Barlow Twins model, a validated SSL framework, to create an unbiased atlas of histomorphologic phenotype clusters (HPCs) from H&E and reticulin-stained BMBx (Fig.1B-1E).^13-15^ These HPCs were reviewed by board-certified hematopathologists to validate clustering patterns, enabling the development of an accurate model distinguishing MDS from clinical mimics.

To assess the model’s generalizability and robustness, 95 unseen MDS cases from MD Anderson Cancer Center (MDACC), Houston, TX, were evaluated using the SSL model.

This study was conducted under the scope of an approved NYU Grossman School of Medicine Institutional Review Board protocol (IRB# i22-00178).

## Results and discussion

### HPCs capture diagnostically meaningful morphologic features

The self-supervised pipeline identified 31 distinct HPCs within the H&E-stained image tiles. Evaluation of the clusters by two hematopathologists validated that most image tiles within each cluster showed similar morphologic features, demonstrating consistent and interpretable feature extraction by the model. HPCs capturing cellular hematopoietic marrow were retained for further analysis and model training. To ensure comprehensive analysis, we incorporated all HPCs into our classifier. However, prior to training, an automated selection step was implemented to identify and prioritize the most relevant HPCs. This approach excluded artifacts and tiles composed entirely of bone tissue without hematopoietic elements, as these were not pertinent to the classification task. Enriched HPCs in MDS cases captured morphologic features relevant for MDS diagnosis, including marrow hypercellularity (HPC #4), dysplastic and clustered megakaryocytes (HPC #4), increased immature hematopoietic cells (HPCs #4, 26), increased vascularity, fibrosis, and cell streaming patterns (HPCs #26).^4,16^ Depleted HPCs in MDS cases were associated with low cellularity (HPCs #9, 21) as well as hematopoietic cells with normal morphologies and maturation (HPCs #6, 12, and 24) (Fig.2A).

**Fig 2.**
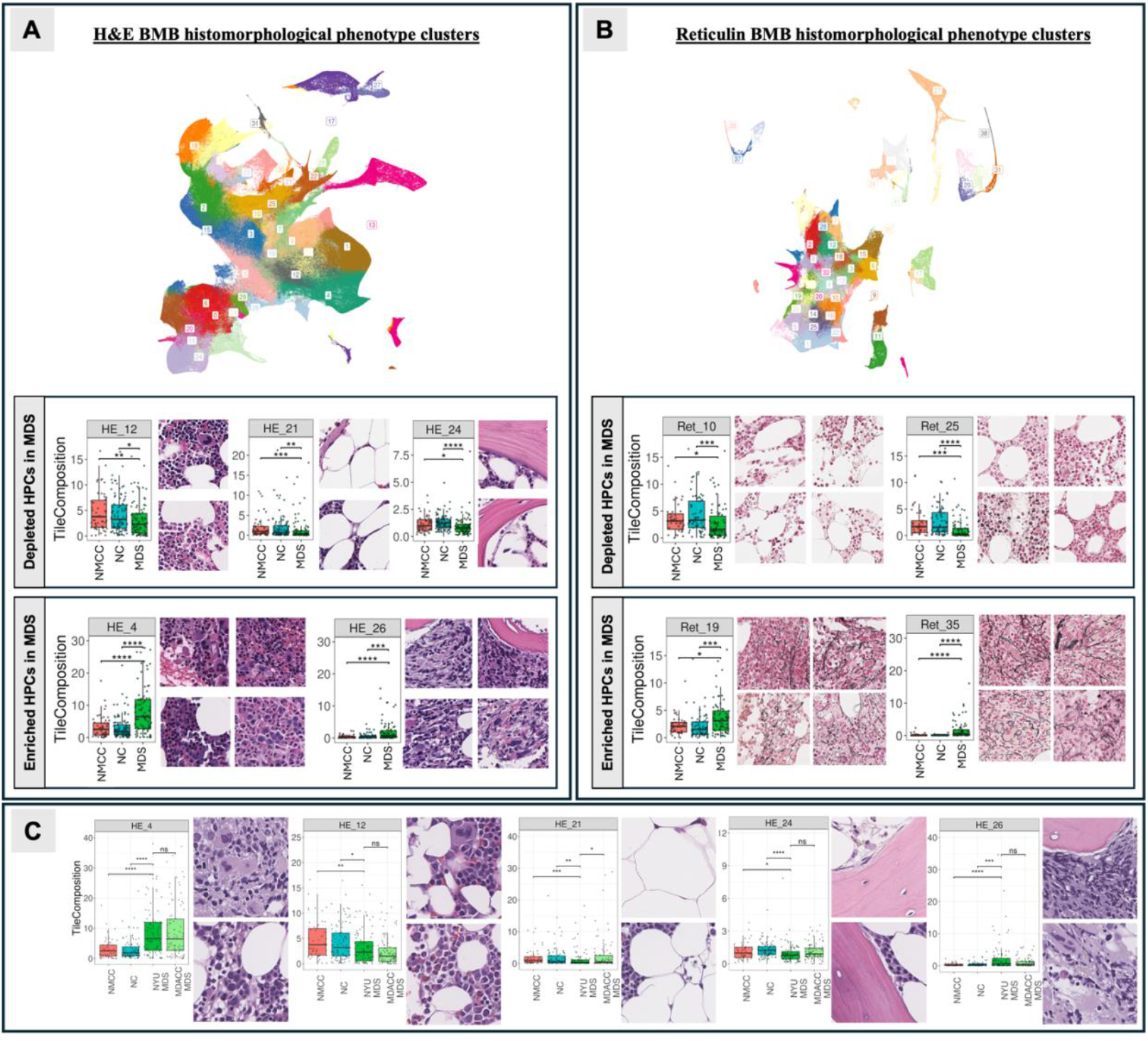
Histomorphological phenotype clusters (HPCs) identified by the model exhibit enrichment patterns in myelodysplastic neoplasms (MDS) that are consistent with known histopathologic features, demonstrating the model's ability to capture diagnostically relevant morphological information; **(A)** Visualization of H&E-stained BMB image tile vector representations using uniform manifold approximation and projection (UMAP) dimensionality reduction, color-coded for HPCs. Representative image tiles from statistically significant enriched and depleted HPCs are depicted, along with their corresponding bar chart (each dot in the bar chart represents the percentage of image tiles belonging to the respective HPC in a single patient); **(B)** Visualization of reticulin-stained BMB image tile vector representations using UMAP dimensionality reduction, color-coded for HPCs. Enriched reticulin HPCs in MDS highlight reticulin fibrosis as the primary feature driving clustering. In contrast, depleted clusters in MDS lack reticulin fibrosis, irrespective of cellularity levels. Representative image tiles from statistically significant enriched and depleted HPCs are depicted; **(C)** External validation with unseen MDS cases from MD Anderson Cancer Center revealed a consistent pattern of HPC enrichment in both cohorts. Image tiles from the independent validation set with similar morphologic features exhibited the same enrichment or depletion patterns observed in the training dataset. [MDS: myelodysplastic neoplasm; NMCC: non-MDS cytopenic control, NC: negative control].

Using the calculated HPC composition for each patient, the model was trained to differentiate between diagnostic groups solely based on H&E WSIs (Fig.3A). The model demonstrated a robust ability to distinguish MDS from NC, yielding an AUC of 0.89. When MDS cases were compared to NMCC cases, the model achieved an AUC of 0.84. Following consolidation of NMCC and NC cases into one group, MDS cases were identified with an AUC of 0.78.

**Fig 3.**
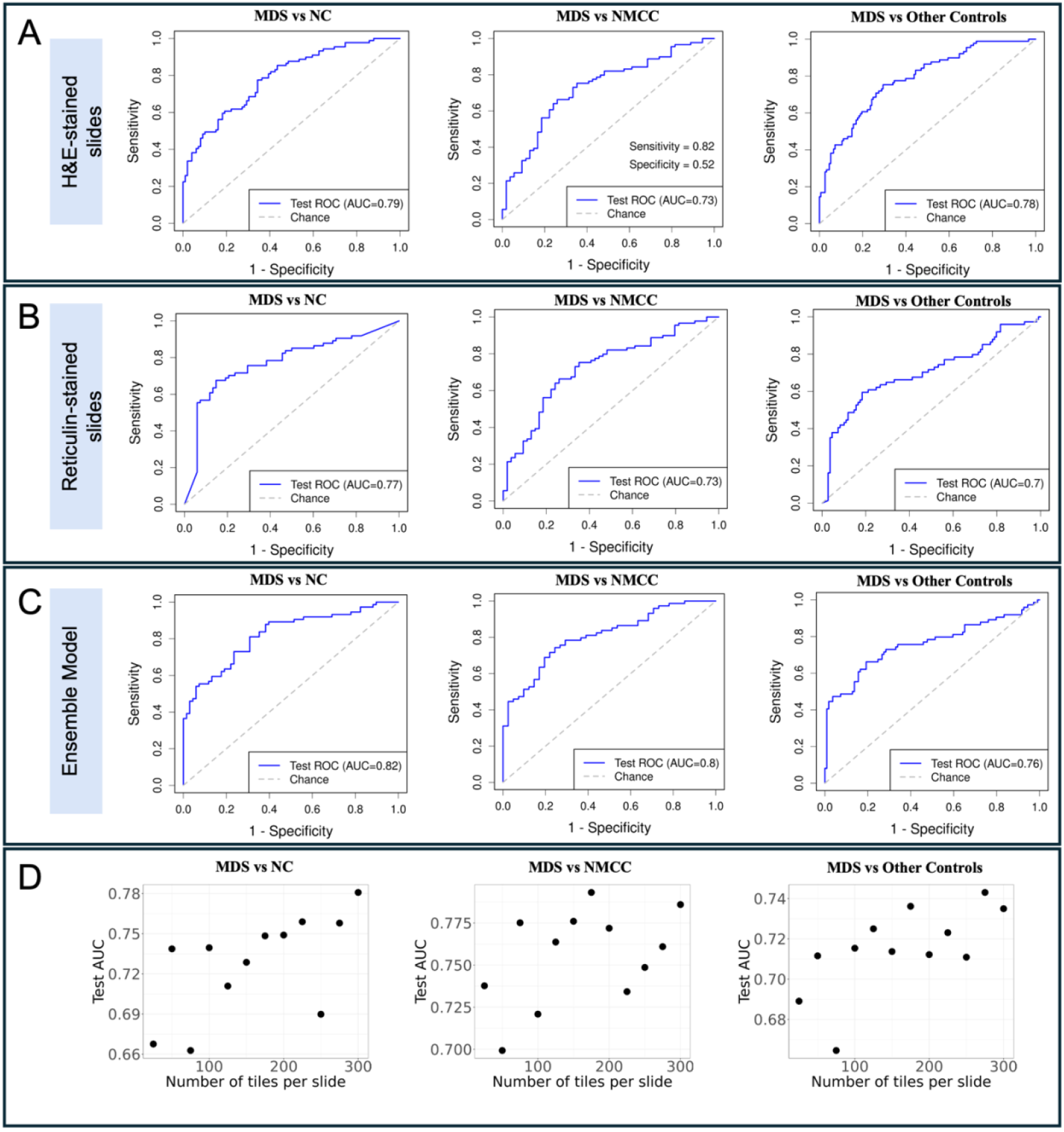
A self-supervised Al image analysis accurately distinguishes MDS from control groups using bone marrow biopsy slides; **(A)** Receiver operating characteristic (ROC) curves for model trained on H&E-stained slides; **(B)** ROC curves for model trained on reticulin-stained slides highlight the influence of reticulin fibrosis on classification; **(C)** ROC curves for the ensemble model, integrating both H&E- and reticulin-derived features, demonstrate improved overall diagnostic accuracy; **(D)** Relationship between the number of image tiles included per slide during training and the resulting test area under the curve (AUC) values for the ensembled model. [MDS: myelodysplastic neoplasm; NMCC: non-MDS cytopenic control, NC: negative control].

A key limitation in diagnosing hematologic disorders is BMBx size, as an adequate amount of intact hematopoietic marrow is essential to accurately represent hematopoiesis.^17^ To determine the minimum amount of tissue required to achieve optimal diagnostic performance, we randomly selected varying numbers of tiles from each WSI. Selecting 200–300 tiles per WSI achieved an AUC comparable to using all tiles (Fig. 3D). Considering a tile size of 224×224 pixels (where 1 pixel = 0.504 μm), the minimum amount of tissue area required to reach optimal performance for this model is approximately 2.55-3.83 mm^2^, which represents significantly less tissue than required for “adequate” samples as defined by current guidelines (1.5 cm in length).^18,19^

### The SSL trained model performs consistently on an external cohort

To evaluate the generalizability of our image analysis algorithm, we assessed the performance on a dataset of BMBx WSIs acquired from an external institution. For this purpose, 95 cases with confirmed diagnosis of MDS from MDACC were analyzed using our trained model. HPCs derived from the NYU dataset were assigned to the MDACC cohort, enabling evaluation of the model’s ability to assign clusters to an independent dataset. Notably, HPCs capturing bone marrow fibrosis and cell streaming (HPC #26), areas of hypercellularity with predominant erythroid hyperplasia (HPC #4), and dysplastic megakaryocytes and immature hematopoietic elements (HPC #4) were enriched in both MDS cohorts. In contrast, image tiles capturing adipocyte enriched areas (HPC #21), normal trilineage maturing hematopoietic cells (HPC #12), and aspiration artifacts (HPC #9) were predominantly enriched in the control groups (Fig. 2C). Indeed, the image tiles of the MDACC MDS cases exhibited similar patterns of cluster enrichment or depletion as observed in the original training dataset derived from NYU cases, highlighting the model’s robustness in recognizing comparable features despite potential staining differences.

### Addition of reticulin stained BMBx to the SSL model increases model performance

One of the features evaluated during routine examination of BMBx for MDS is the degree of reticulin fibrosis, with diagnostic and prognostic implications.^4,20,21^ We analyzed WSIs of reticulin-stained BMBx using the same self-supervised pipeline.

Analysis of WSIs of reticulin-stained BMBx identified 39 HPCs with a subset enriched in MDS cases. Evaluation of images within each HPC revealed that the clusters and enrichment patterns were primarily driven by the presence and density of reticulin fibrosis. Although morphological features identified in H&E-stained slides were still discernible in reticulin-stained slides due to the nuclear counterstain, the clustering of image tiles appeared to be primarily driven by features related to fibrosis, since HPCs in reticulin-stained slides exhibiting these cellular features did not influence the clustering as expected (Fig.2B). The trained model using only the reticulin-stained slides distinguished between MDS vs NC, and MDS vs NMCC with AUCs of 0.85, and 0.81, respectively.

Subsequently, an ensemble model was trained to integrate the results from both reticulin- and H&E-stained WSIs. The ensemble model improved the classification performance, reaching AUCs of 0.89, 0.84, and 0.78 in distinguishing between MDS vs NC, MDS vs NMCC, and MDS vs other controls together, respectively (Fig.3C).

Notably, we tested multiple conditions to optimize the model. We varied tile image sizes based on our prediction that smaller tiles (50x50 and 12×12 pixels) capturing fewer cells might produce models that focus on cytologic features and thus might be more robust in identifying dysplastic cells. However, our most robust model was based on the 224×224 pixel (113μm/113μm) tiles, suggesting that changes in marrow cell composition and spatial relationships are features of MDS that distinguish it from other acquired causes of cytopenia. This finding is consistent with prior studies that showed that disruption of erythroid islands or other alterations in cell composition are characteristic of MDS.^22^

A limitation of our study is the lack of genetic and mutation information on all the NMCC; therefore, we were unable to reliably annotate and distinguish cases of clonal cytopenia of uncertain significance (CCUS) from idiopathic cytopenia of uncertain significance. We envision that studying larger, genetically annotated CCUS cohorts alongside MDS will reveal early, subtle MDS features, facilitating earlier diagnosis of disease.

While the results suggest that SSL models may be deployable in the clinical diagnostic setting, ultimate adoption will require validation using a larger number of cases from additional institutions. We envision such models will not be free-standing but will be integrated with pathologist-trained models to improve accuracy since features that pathologists routinely evaluate such as megakaryocyte dysplasia were not featured prominently among the HPCs enriched in MDS. We anticipate that integrated models will benefit from immunohistochemical stains (e.g. CD34, CD61, GPA). Such integrated approaches can be tested for their ability to assess mutational subtypes or clinical outcomes.

## Data Availability

All data produced in the present study are available upon reasonable request to the authors.

## Acknowledgments

The authors acknowledge the contributions of the Experimental Pathology Research Laboratory, particularly Cynthia Loomis and Valeria Mezzano, and the Center for Biospecimen Research & Development, particularly Andre L. Moreira, Luis A. Chiriboga, Faye Bourie, and Itana Rekhelis.

## Authorship

Contribution: Conceptualization: AT, CYP; Data Acquisition: VM, BJ, SL, GGM; Data Analysis: VM, HL, BJ, SL, AT, CYP; Writing – Original Draft: VM, HL, CP; Manuscript Review & Editing: VM, HL, BJ, SL, AT, CP, GGM.

## Conflict-of-interest disclosure

SL: Consultancy/Honoraria: Recordati, Daiichi-Sankyo, AlphaSights, Gerson Lehrman Group, Arima Genomics, Ground Truth Labs, Qiagen, Servier, Syndax, Kura, AbbVie, Cogent, Tempus AI, Caris Diagnostics; Stock Ownership: AbbVie.

## Notes

### Funding Statement

This study was funded by NIH/NCI (R01CA249054).

### Author Declarations

Ethics committee/IRB of NYU Grossman School of Medicine gave ethical approval for this work (IRB# i22-00178).

