## Supplemental file for "Self-Supervised Learning Can Distinguish Myelodysplastic Neoplasms from Clinical Mimics Using Bone Marrow Biopsies"

### Methods:

#### Patient Selection and Control Group Assignment

A total of 243 BMB's were included in the study, representing 89 diagnostic myelodysplastic neoplasm (MDS), 55 cytopenic, non-MDS controls ("non-MDS cytopenic controls" [NMCC]), and 99 negative control cases ("negative control" [NC]). MDS cases were selected based on established diagnostic criteria, including clinical cytopenia, bone marrow histopathologic evaluation, cytogenetic analysis, and molecular studies. Among these, 26 patients were diagnosed with excess blasts-2, 18 with excess blasts-1, and 45 with other MDS subtypes. Lymphoma staging marrow cases were included as NC if they had confirmed lymphoma diagnoses and bone marrow biopsies showing no dysplastic features or any other morphological abnormalities. Cytopenic cases undergoing BMBx procedures with suspicion of MDS but resulting in non-MDS diagnoses were selected as more challenging controls. These cases encompassed a range of conditions, including autoimmune disorders, solid organ tumors, alcohol-related conditions, and chronic liver and kidney disorders, all confirmed through thorough clinical, laboratory, and histopathological evaluations. For the NC, cases were included that had negative results on the NYU Oncomine Myeloid panel, a multi-biomarker NGS assay that detects variants in 50 key genes related to hematological malignancies, indicating the absence of mutations associated with myeloid malignancies.[1,2] Cases with poor-quality BMBx or inadequate cellularity (less than one intertrabecular space) were excluded to ensure accurate interpretation. Additionally, any cases with inconclusive diagnoses were excluded from the study.

### Self-Supervised Learning Image Analysis Pipeline

An SSL framework, Barlow Twins model, previously validated across various tissue types, was employed to construct a comprehensive and unbiased atlas of histomorphologic phenotype clusters (HPCs) derived from hematoxylin and eosin H&E and reticulin-stained specimens.[3] This atlas was subsequently utilized to train a classifier for disease categorization based on distinct HPC profiles by comparing confirmed MDS BMBx to those from NMCC and NC.[4-6] Each HPC was examined by hematopathologists, resulting in a detailed examination of morphologic features, enabling the validation of clustering patterns. This resulted in the development of a highly accurate model that distinguished MDS from age-matched, cytopenic, non-MDS controls. Overall, these studies demonstrate the utility of SSL approaches to identify morphologic features that allow reproducible and accurate discrimination of MDS from its clinical mimics.

### Whole Slide Image Preprocessing

We used the publicly available pipeline DeepPATH to segment each WSI into non-overlapping 224 x 224 tiles at 20X magnification (0.504µm/pixel).[6] Tiles with 100% background pixels were excluded. Background pixels were defined as pixels with a grey level above 230. We applied Reinhard's method for stain normalization.[7] We split all patients into balanced training (80%) and validation (20%) sets.

### Overview of Histomorphological Analysis and Classification Pipeline

All available tissue sections, typically two slides per patient with two levels per slide, were included in the analysis to increase sampling depth, improve representativeness, and mitigate sectional variability, by covering more tissue section levels.

Whole slide images (WSIs) of H&E-stained BMB sections were obtained at 40x and segmented as 224x224 pixel tiles at 20x magnification to ensure high-resolution capture of tissue morphologies. This resulted in 302 to 18967 tiles per case with an average of 3464 tiles per case. The tiled images underwent a stain normalization process to standardize color variations, facilitating more accurate comparisons across samples and mitigating the impact of variations in the imaging process across different institutions.[6,7] The preprocessed tiles were input into a self-supervised learning framework, the Histomorphological Phenotype Learning pipeline (HPL)[5], which has proven to be effective in capturing complex histomorphological features in lung adenocarcinoma as well as in pan-cancer datasets. An SSL model was trained on all 1,263,041 tiles from the training dataset then each of the 1,564,617 tiles from our complete dataset were projected into the frozen trained network. The self-supervised pipeline resulted in a 128-dimensional latent space representation for each image tile, effectively encoding complex morphological features into a structured, high-dimensional vector space.

The high-dimensional latent space representations were subjected to dimensionality reduction using Uniform Manifold Approximation and Projection (UMAP).[8] Histomorphological phenotype clusters (HPCs) were then identified using the Leiden community detection algorithm with an unsupervised resolution selection, which operated on reduced-dimensional data to detect meaningful clusters based on histomorphologic features. For each patient, the composition of these HPCs was calculated, resulting in a vectorized representation that reflected the frequency and distribution of the clusters within their tissue samples. These cluster compositions were then used to train a logistic regression model aimed at predicting the disease group based on the identified HPCs (Fig 1B-E).

**Histomorphological Phenotype Learning (HPL) Training**

Of these cases, 74 MDS, 42 NMCC, and 68 NC also had reticulin-stained slides available for image analysis. All available tissue sections, typically two slides per patient with two levels per slide, were included in the analysis to increase sampling depth, improve representativeness, and mitigate sectional variability, by covering more tissue section levels.

We trained each image modality separately for the self-supervised task. Our training set was composed of 1,263,041 tiles and our validation set of 301,576 tiles. We trained a Barlow Twins model for 60 epochs with a batch size of 64.[5] We projected all tiles into the frozen trained networks and obtained tile vector representations of dimensionality 128.

We set up a new balanced training (80%), validation (10%) and test (10%) sets for all patients. Using K nearest neighbors and Leiden community detection on the training set, we identified 32 clusters (HPCs) from the H&E tile vector representations and 52 HPCs from the reticulin tile vector representations. We defined separate H&E and reticulin patient HPC representations with dimensionality equal to the number of HPCs and with each dimension defined as the percentage contribution of a HPC to the total number of tiles from a patient.

**Evaluation of MDS HP-Atlas**

To evaluate the H&E HPCs, we preprocessed the H&E WSIs from our MDACC cohort, projected its tiles into the frozen train H&E network and assigned HPCs to the resulting tile vector representations. We defined patient HPC representations and compared them to our internal NYU patient HPC representations with Wilcoxon tests. Additionally, we asked an expert to evaluate the consistency of the clusters in the MDACC cohort to make sure the same patterns were represented across both cohorts.

### MDS Classification Model

We employed a balanced 5-fold cross-validation approach, stratified by MDS subtypes and control groups, to train logistic regression models using H&E patient HPC representations, reticulin patient HPC representations, and their integration for predicting MDS versus control groups. Cluster selection was automated on each fold train set using the Wilcoxon test to identify significant clusters ($p \leq 0.05$). All patient HPC representations were normalized with centered-log ratio (CLR) and only selected HPCs were used for prediction in each fold. For the model integrating both image modalities, we trained an ensemble model with weights optimized through grid search to maximize the area under the curve (AUC) on the train set, combining predictions from the individual models based on H&E patient HPC representations and reticulin patient HPC representations. The AUC values displayed in the figures represent the performance calculated by pooling the predictions from all 5 folds, with a single AUC computed for the combined test set predictions.

### Robustness of the MDS Classification Model

To evaluate the robustness of our MDS classification models, we conducted additional experiments using a reduced number of tiles per patient. We randomly selected 25, 50, 75, 100, 125, 150, 175, 200, 225, 250, 275 and 300 contiguous tiles per patient, recomputed all patient HPC representations, and retrained all our classifiers to test whether the model could maintain strong predictive performance despite having less data to work with.

**External H&E Validation Dataset:** To evaluate our self-supervised findings, we used an independent external MDS cohort from MD Anderson Cancer Center (MDACC). This dataset was composed of 95 H&E-stained whole slide images (WSIs) from 95 MDS patients.
